# Real-time heart rate variability according to ambulatory glucose profile in patients with diabetes mellitus

**DOI:** 10.1101/2023.05.04.23289543

**Authors:** Sung Il Im, Su Hyun Bae, Soo Jin Kim, Bong Joon Kim, Jung Ho Heo, Su kyoung Kwon, Sung Pil Cho, Hun Shim, Jung Hwan Park, Hyun Su Kim, Chul Ho Oak

**Affiliations:** Division of Cardiology, Department of Internal Medicine, Kosin University Gospel Hospital, Kosin University College of Medicine, Busan, Korea; Division of Endocrinology, Department of Internal Medicine, Kosin University Gospel Hospital, Kosin University College of Medicine, Busan, Korea; Me-zoo, Won Ju, Gangwon province, Korea; Division of Pulmonology, Department of Internal Medicine, Kosin University Gospel Hospital, Kosin University College of Medicine, Busan, Korea

**Keywords:** heart rate variability, glucose level, real-time monitoring

## Abstract

**Background:** Autonomic neuropathy commonly occurs as a long-term complication of diabetes mellitus (DM) and can be diagnosed based on heart rate variability (HRV), calculated from electrocardiogram (ECG) recordings. There are limited data on HRV using real-time ECG and ambulatory glucose monitoring in patients with DM.

**Methods and Results:** A total of 43 patients (66.3±7.5 years) with DM underwent continuous real-time ECG monitoring (225.7±107.3 hours) for HRV and ambulatory glucose monitoring using a remote monitoring system. We compared the HRV according to the ambulatory glucose profile. Data were analyzed according to the target in glucose range (TIR). There were no significant differences in the baseline characteristics of the patients according to the TIR. During monitoring, we checked ECG and ambulatory glucose levels (a total of 15,090 times) simultaneously for all patients. Both time- and frequency-domain HRVs were lower when the patients had poorly controlled glucose levels (>200 mg/dL) and TIR<70% compared with normally controlled glucose levels (<200 mg/dL) and TIR>70%. In addition, heart and respiratory rates increased with real-time glucose levels (P<0.001).

**Conclusions:** Poorly controlled glucose levels were independently associated with lower HRV in patients with DM. This was further substantiated by the independent continuous association between real-time measurements of hyperglycemia and lower HRV. These data strongly suggest that cardiac autonomic dysfunction is caused by elevated blood sugar levels.

## Introduction

Heart rate variability (HRV) is the fluctuation in the time interval between adjacent heartbeats.^1^ Cardiac autonomic function can be noninvasively assessed by calculating HRV, which reflects the interaction of the sympathetic and parasympathetic parts of the autonomic nervous system (ANS) on the sinus node. HRV indexes neurocardiac function and is generated by heart-brain interactions and dynamic nonlinear ANS processes. HRV is an emergent property of the interdependent regulatory systems that operate at different timescales to help us adapt to environmental and psychological challenges. HRV reflects the regulation of autonomic balance, blood pressure (BP), gas exchange, and gut, heart, and vascular tone, which refers to the diameter of the blood vessels that regulates BP.^2^

Type 2 diabetes mellitus (DM) is increasingly prevalent worldwide and is associated with an increase in obesity and metabolic syndrome.^3^ The number of people with DM is predicted to double within the next three decades.^4^ Besides macrovascular and microvascular complications, the leading causes of death in DM are cardiovascular complications. Cardiovascular mortality is associated with cardiac autonomic neuropathy, which is frequently associated with DM.^5^

Screening for cardiac autonomic neuropathy is recommended for the diagnosis of DM, particularly in patients with a history of poor glycemic control, macro and microvascular complications, and increased cardiovascular risk. Although standard cardiovascular reflex tests remain the gold standard for the assessment of cardiovascular autonomic neuropathy, one of the easiest and most reliable ways to assess cardiac autonomic neuropathy is by measuring HRV. HRV is the variation between two consecutive beats; the higher the variation, the higher the parasympathetic activity.^6^ A high HRV reflects the fact that an individual can constantly adapt to microenvironmental changes. Therefore, low HRV is a marker of cardiovascular risk.^7^ Conveniently, the measurement of HRV is non-intrusive and pain-free.^1^

Although the evaluation of HRV in DM has been assessed in several studies, conflicting results have been reported.^2,3,6^ Moreover, there is no consensus on the decreased levels of HRV parameters in patients with DM. Furthermore, despite the link between HRV and DM severity,^8^ there are limited data on the association between HRV parameters and glucose levels using real-time electrocardiogram (ECG) and ambulatory glucose monitoring in patients with DM. Therefore, we aimed to simultaneously check HRV and glucose levels in patients with DM to identify the most explanatory variables for autonomic dysfunction according to the glucose level.

## Methods

### Participants

We recruited 43 patients (mean age, 65.5±6.2 years) with DM from endocrinology out-patient clinic during their usual follow-up. The participants were recruited between October 2021 and December 2021. All patients were screened for medication use and medical conditions.

The inclusion criteria were age>18 years, type 2 DM, and treatment with oral antidiabetic agents. The main exclusion criteria were pregnancy, neurological disease, heart failure, chronic liver or renal failure (known chronic liver disease or stage 3 advanced chronic kidney disease), uncontrolled DM, thyroid disorder, or treatment that could influence HRV parameters.

In our study, normal candidates (40 patients) without DM were included as controls. Five patients who were lost to follow-up or had incomplete monitoring were excluded from the study. Before HRV measurements, patients answered a questionnaire on personal information and lifestyle habits (e.g., smoking, alcohol consumption, coffee drinking, and exercise).

Finally, total 38 patients (16 men and 22 women; mean age: 66.3±7.5 years) who completed the HRV measurements and glucose monitoring were included in the analysis.

The study protocol was approved by the ethical guidelines of the 1975 Declaration of Helsinki, and the research protocol (IRB No. 2022-06-016) was approved by the Ethics Committee of Kosin University Gospel Hospital. Written informed consent was obtained from all patients.

### Data collection

After ECG and chest radiography, the cardiovascular status of each patient was evaluated using echocardiography and blood laboratory data from the initial visit, as determined by the attending physicians. From the database, the following information were collected: (1) patient data, including sex, age, height, and weight; (2) cardiovascular risk factors, including hypertension (use of antihypertensive agents, systolic blood pressure≥140 mmHg, or diastolic blood pressure≥90 mmHg on admission) and DM (use of oral hypoglycemic agents or insulin, or glycosylated hemoglobin≥6.5%); (3) cardiovascular disease status, including structural heart disease, congestive heart failure, or a history of a disabling cerebral infarction or transient ischemic attack; and (4) use of medication.

### ECG monitoring device

Hicardi® (MEZOO Co., Ltd., Wonju-si, Gangwon-do, Korea) is an 8 g, 42×30×7 mm (without disposable electrodes) wearable ECG monitoring patch device certified as a medical device by the Ministry of Food and Drug Safety of Korea (supplement figure 1). This wearable device monitors and records single-lead ECGs, respiration, skin surface temperature, and activity. The ECG signal is recorded with a 250 Hz sampling frequency and 14-bit resolution.

The data from the wearable patch were transferred through Bluetooth Low Energy to a mobile gateway, which was implemented as a smartphone application. The mobile gateway transmitted the data to a cloud-based monitoring server.

After informed consent was obtained from the patient, a wearable patch was attached to the left sternal border. The ECG signals and the above-mentioned data were continuously recorded, and all ECG signals were reviewed by a cardiologist via a cloud-based monitoring server.

### HRV parameters

HRV analysis was performed in the time and frequency domains of the wearable ECG recordings according to international guidelines [29].

On average, 225.7±107.3 h of ECG were recorded per patient, and the HRV analysis was performed by excerpting the previous five-minute segment from the time of glucose measurement.

To calculate the HRV parameters, RR intervals must be computed from the wearable ECG recordings. The following steps were performed to obtain the RR interval time series. First, R-peaks were detected using the geometric angle between two consecutive samples of the ECG signal.^9^ Detected R-peaks were then used to generate an RR interval time series. To remove the abnormal intervals caused by ectopic beats, arrhythmic events, missing data, and noise, intervals below 80% or above 120% of the average of the last six intervals were excluded. The time-domain parameters were calculated from the RR interval time series. Second, the RR interval time series was resampled at 4 Hz using linear interpolation. The resulting series was detrended by eliminating linear trends. After detrending, the power spectral density for the RR interval time series was estimated using the Burg autoregressive model, where the order of the model was 33.

In the time domain, we analyzed the RR intervals, standard deviations of RR intervals, square root of the mean squared difference of successive RR intervals, and percentage of adjacent NN intervals differing by more than 50 ms (NN50).

In the frequency domain, we analyzed low frequency (LF, 0.04–0.15 Hz), an index of both sympathetic and parasympathetic activity, and high frequency (HF, 0.15–0.4 Hz), representing the most efferent vagal (parasympathetic) activity to the sinus node. Very low frequency (VLF; 0.003–0.04 Hz) partially reflects thermoregulatory mechanisms, fluctuations in the activity of the renin–angiotensin system, and the function of peripheral chemoreceptors. The LF/HF ratio, that is, sympathovagal balance, was also calculated.

### Continuous glucose monitoring

#### Assessment of glucose status

For continuous glucose monitoring, we used FreeStyle Libre 14 day system®, a continuous glucose monitoring device with real-time alarm capability indicated for the management of DM. The flash glucose-sensing technology used was the FreeStyle LibreTM, which is a sensor-based flash glucose-monitoring system (Abbott Diabetes Care, Witney, UK). The sensor was worn on the back of the arm for up to 14 days, and glucose data were automatically stored every 15 min. Real-time glucose levels can be obtained as often as every minute by scanning the sensor with a reader. Data were transferred wirelessly by radio-frequency identification from the sensor to the reader’s memory, which stored historical sensor data for 90 days. Data can be uploaded using the device software to generate summary glucose reports. The target in glucose range (TIR) was 70-180 mg/dL. We analyzed the data according to glucose control and TIR of <70% or >70%. For these individuals, fasting glucose levels and information about DM medication were used to determine glucose metabolism status. Glucose metabolism status was defined according to the 2006 World Health Organization criteria as normal glucose metabolism or type 2 diabetes.^10^

### Statistical analysis

All continuous variables are expressed as mean±standard deviation (SD) or median (25th and 75th interquartile range), depending on the distribution. For continuous data, statistical differences were evaluated using the Student’s *t-*test or Mann–Whitney *U* test, depending on the data distribution. Categorical variables are presented as frequencies (percentages) and were analyzed using the chi-squared test. To determine whether any of the variables were independently related to HRV according to the glucose levels, a multivariate analysis of variables with a P-value<0.05 in the univariate analysis was performed using linear logistic regression analysis. All correlations were calculated using the Spearman’s rank correlation test. All statistical analyses were conducted using the SPSS statistical software (version 19.0 (SPSS Inc., Chicago, IL, USA), and statistical significance was set at P <0.05 (two-sided).

## Results

A total of 38 patients (age, 66.3±7.5 years) with DM underwent continuous real-time ECG monitoring (225.7±107.3 hours) for HRV and ambulatory glucose monitoring using a remote monitoring system. We compared the HRV according to the ambulatory glucose profile. Ambulatory glucose levels were checked every 15 minutes in all patients during real-time ECG monitoring.

During monitoring, we checked a total of 15,090 ECG data points for HRV and ambulatory glucose levels simultaneously for all patients. We analyzed the data according to the TIR. There were no significant baseline differences in patient characteristics except for the mean glucose level according to the TIR (Table 1). No significant difference in baseline medication, according to the TIR, was observed (Table 2).

**Table 1.** Baseline characteristics according to TIR in patients with DM.

| Total patients = 38 |  |  |  |
| --- | --- | --- | --- |
| Variable | TIR<70 % (n=13) | TIR>70 % (n=25) | P value |
| Mean glucose level (mg/dL) | 241.1±60.3 | 129.3±27.3 | <0.001 |
| Age (years) | 64.3±6.2 | 66.2±6.1 | 0.161 |
| Sex (%), male | 6 (46.2) | 14 (56.0) | 0.734 |
| DM (%) | 13 (100) | 25 (100) | 1.000 |
| HTN (%) | 12 (92.3) | 18 (72.0) | 0.222 |
| Hyperlipidemia (%) | 11 (84.6) | 19 (76.0) | 0.689 |
| CAD (%) | 3 (23.0) | 9 (36.0) | 0.486 |
| CVA (%) | 2 (15.4) | 6 (24.0) | 0.689 |
| CHF (%) | 0 (0) | 2 (12.0) | 0.538 |
| CMP (%) | 1 (7.7) | 4 (16.7) | 0.638 |

\*TIR indicates target in glucose range; DM, diabetes mellitus; HTN, hypertension; CAD, coronary artery disease; CVA, cerebrovascular accident; CHF, congestive heart failure; CMP,
cardiomyopathy.

**Table 2.**
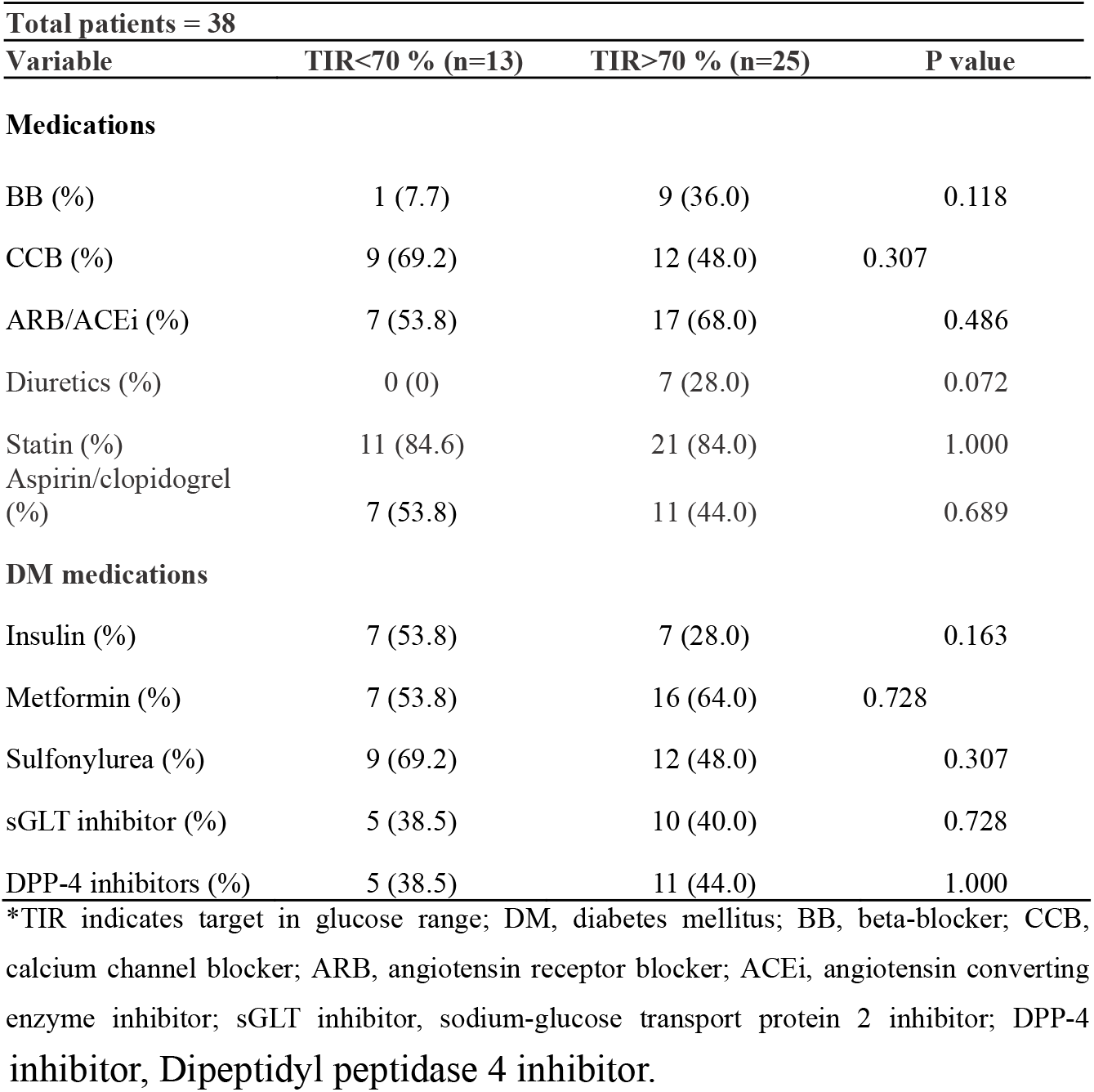
Baseline medicafions according to TIR in patients with DM.

| **Total patients = 38** |  |  |  |
| --- | --- | --- | --- |
| **Variable** | **TIR<70 % (n=13)** | **TIR>70 % (n=25)** | **P value** |
| **Medications** |  |  |  |
| BB (%) | 1 (7.7) | 9 (36.0) | 0.118 |
| CCB (%) | 9 (69.2) | 12 (48.0) | 0.307 |
| ARB/ACEi (%) | 7 (53.8) | 17 (68.0) | 0.486 |
| Diuretics (%) | 0 (0) | 7 (28.0) | 0.072 |
| Statin (%) | 11 (84.6) | 21 (84.0) | 1.000 |
| Aspirin/clopidogrel (%) | 7 (53.8) | 11 (44.0) | 0.689 |
| **DM medications** |  |  |  |
| Insulin (%) | 7 (53.8) | 7 (28.0) | 0.163 |
| Metformin (%) | 7 (53.8) | 16 (64.0) | 0.728 |
| Sulfonylurea (%) | 9 (69.2) | 12 (48.0) | 0.307 |
| sGLT inhibitor (%) | 5 (38.5) | 10 (40.0) | 0.728 |
| DPP-4 inhibitors (%) | 5 (38.5) | 11 (44.0) | 1.000 |

\*TIR indicates target in glucose range; DM, diabetes mellitus; BB, beta-blocker; CCB, calcium channel blocker; ARB, angiotensin receptor blocker; ACEi, angiotensin converting enzyme inhibitor; sGLT inhibitor, sodium-glucose transport protein 2 inhibitor; DPP-4
inhibitor, Dipeptidyl peptidase 4 inhibitor.

Both time- and frequency-domain HRVs were lower in patients with poorly controlled glucose levels (TIR<70%) than in those with normally controlled glucose levels (TIR>70%; Table 3).

**Table 3.** HRV measures according to TIR in patients with DM.

| **HRV** |  |  |  |
| --- | --- | --- | --- |
| **Time domain** | **TIR<70%** | **TIR>70%** | **P value** |
| SDNN, ms | 40.5 ± 38.5 | 48.1 ± 43.1 | <0.001 |
| RMSSD, ms | 7.9 ± 6.9 | 10.0 ± 9.2 | <0.001 |
| SDSD, ms | 7.9 ± 6.9 | 10.1 ± 9.2 | <0.001 |
| NN50, count | 14.6 ± 40.7 | 25.3 ± 55.6 | <0.001 |
| pNN50, % | 1.0 ± 2.9 | 1.8 ± 4.0 | <0.001 |
| **Frequency domain** |  |  |  |
| Total Power, N.U. *10<sup>5</sup> | 5.5 ± 0.3 | 8.2 ± 0.3 | <0.001 |
| VLF, N.U. *10<sup>5</sup> | 3.7 ± 0.3 | 4.7 ± 0.2 | 0.019 |
| LF, N.U. *10<sup>5</sup> | 1.2 ± 0.3 | 2.3 ± 0.5 | <0.001 |
| HF, N.U. *10<sup>5</sup> | 0.6 ± 0.1 | 1.3 ± 0.3 | <0.001 |

\*HRV indicates heart rate variability; TIR, target in glucose range; DM, diabetes mellitus; SDNN, standard deviation of NN intervals; RMSSD, root mean square of successive RR interval differences; SDSD, standard deviation of differences between adjacent NN intervals; NN50, number of NN intervals differed by more than 50ms; pNN50, ratio of NN50; Total power, 5 min total power in frequency range ≤0.4 Hz; VLF, power in very low frequency range ≤0.04 Hz; LF, power in low frequency range 0.04 - 0.15 Hz; HF, power in high frequency range 0.15 - 0.4 Hz; N.U., normalized unit.

In addition, heart and respiratory rates increased according to real-time glucose levels (P<0.001) in all patients with DM (Figure 1).

**Figure 1.**
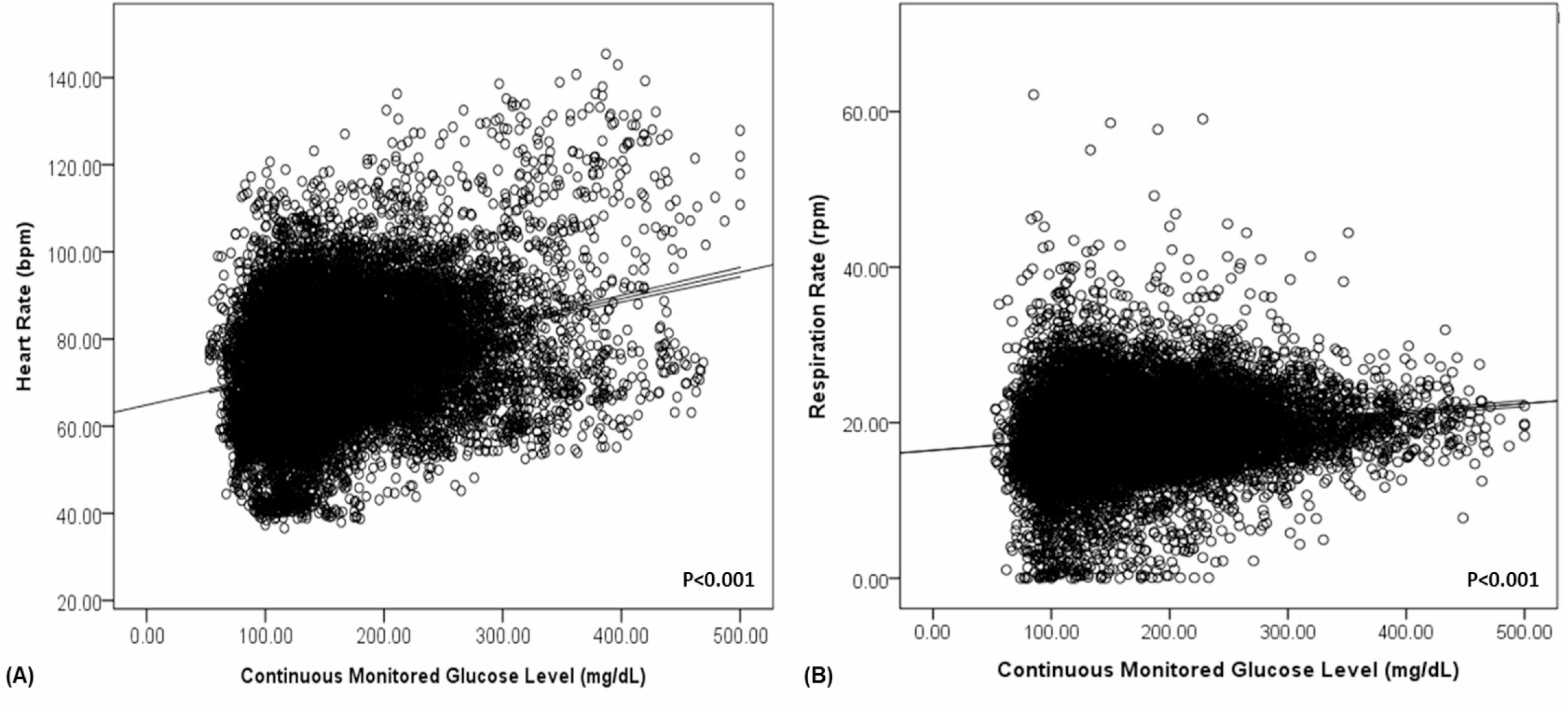
Heart rate (A) and respiration rate (B) according to continuously monitored glucose levels in patients with DM.

As shown in Figure 2, continuous measures of glycemia (plasma glucose levels) were linearly associated with HRV (time domain, NN50 (A); frequency domain, HF (B); P<0.001). As shown in Figure 3A, we compared the frequency domain of HRV (LF) according to DM and TIR. The patients with DM had a lower LF than those without DM (P<0.001). Patients with DM and TIR<70% also had a lower LF than those with TIR>70% (P<0.001).

**Figure 2.**
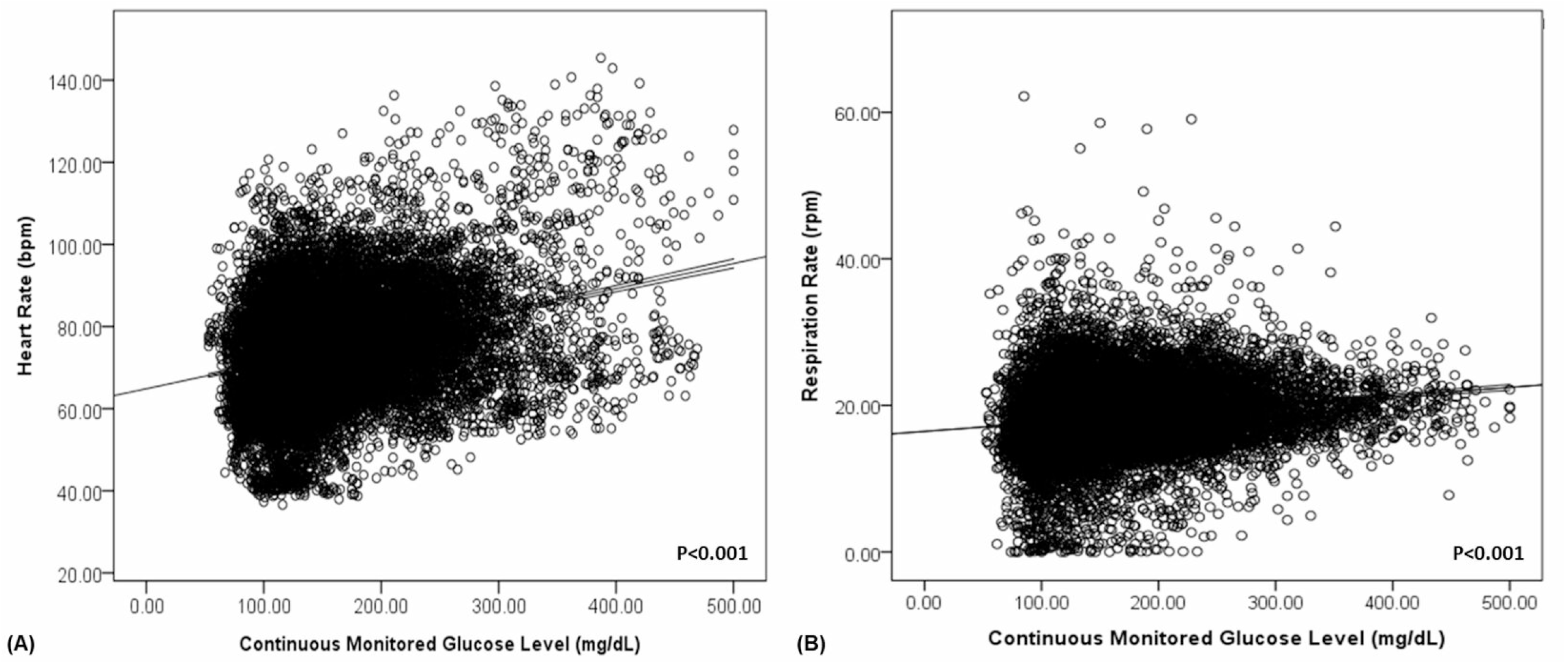
Heart rate variability (HRV) according to continuously monitored glucose level in the time domain [NN50, (A)] and frequency domain [HF, (B)]

**Figure 3.**
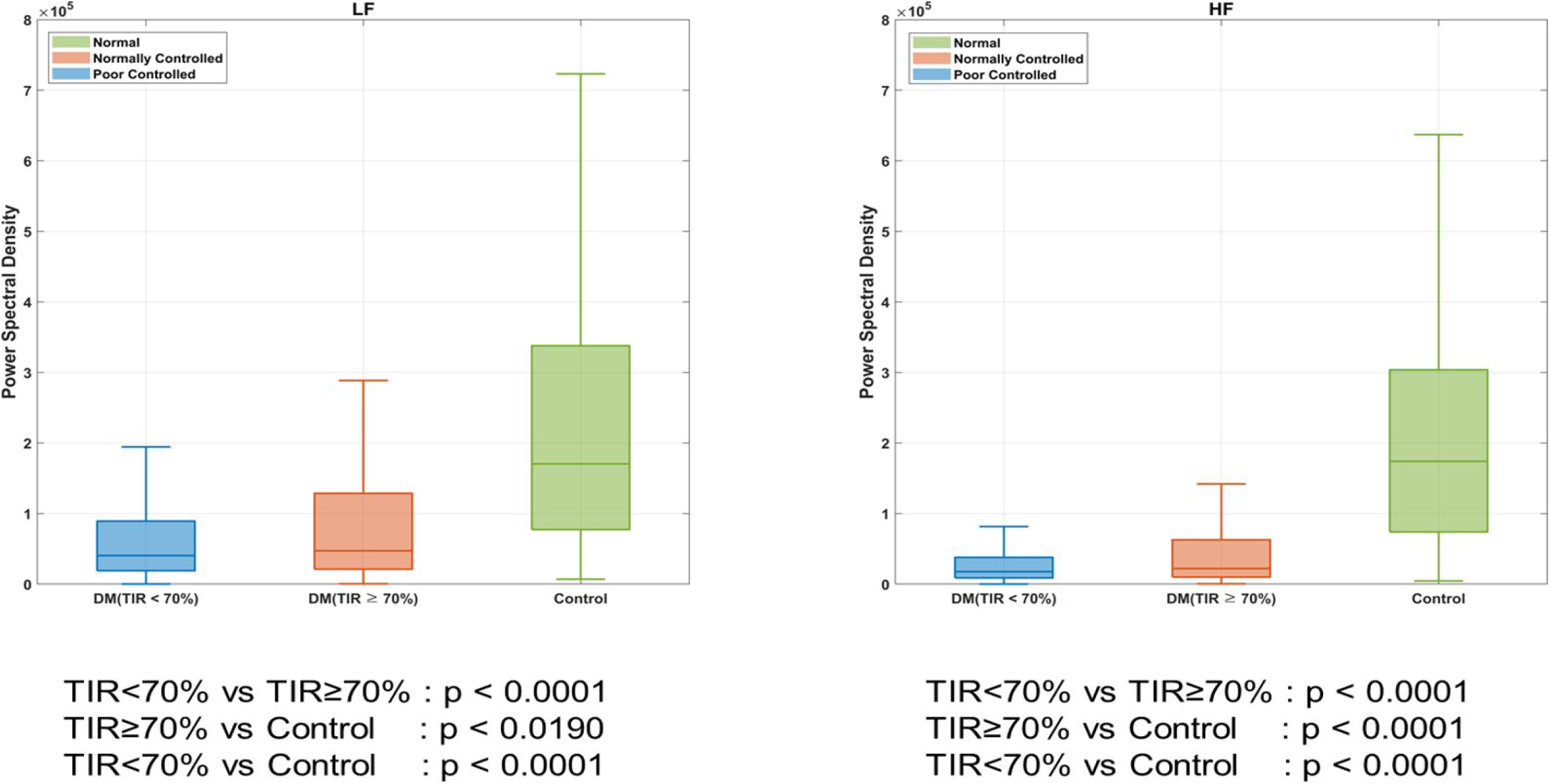
Heart rate variability (HRV) according to DM. *DM indicates diabetes mellitus; TIR, target in glucose range.

As shown in Figure 3B, we compared the frequency-domain HRV (HF) according to DM and TIR. The patients with DM had a lower HF than those without DM (P<0.001). Patients with DM and TIR<70% had a lower HF than those with TIR>70% (P<0.001).

In our study, we found that patients with DM and TIR<70% had a lower HRV than those with TIR>70%, and all patients with DM had a lower HRV than those without DM.

## Discussion

In this study, we simultaneously evaluated heart rate and HRV according to glucose levels in patients with DM. The results of the current study demonstrated that poorly controlled DM is associated with lower HRV. The amount by which HRV was lower in patients with DM with TIR<70% (compared to those with TIR>70%) was approximately 2/3 in both the time and frequency domains. In addition, continuous measures of glycemia (plasma glucose levels) were linearly associated with HRV, suggesting a graded decline in HRV with worsening glucose tolerance. Heart and respiratory rates increased according to real-time glucose levels in all patients with DM. These associations were independent of the major cardiovascular risk factors. Therefore, our results support the concept that cardiac autonomic dysfunction occurs when poorly controlled glucose levels are measured in real time before checking long-term glucose level predictors such as HbA1c and C-peptide levels and may play a role in the development of cardiovascular diseases earlier in the course of type 2 DM.

Cardiac autonomic dysfunction is a complication of DM that carries an approximately fivefold increased risk of mortality in adults.^2^ Damage to the autonomic innervations of the heart and blood vessels can lead to lethal arrhythmias and sudden cardiac death.^11^ Hyperglycemia is thought to be associated with abnormal signaling of autonomic neurons via accumulation of advanced glycation end products, activation of polyol pathway, and ischemia induced atrophy of the autonomic nerve fibers innervating the cardiac and vascular tissues.^12^ Both divisions of the ANS are typically affected, with parasympathetic impairment preceding the sympathetic dysfunction.^6^ Loss of HRV is one of the earliest manifestations of this process. In the Framingham Heart Study, HRV was found to be inversely associated with the risk of mortality.^13^ Similarly, the Atherosclerosis Risk in Communities study found that decreased HRV was independently associated with the risk of developing coronary heart disease.^14^ and lower HRV was also associated with the total burden of cerebral small vessel disease (CSVD) and each of the magnetic resonance image markers of CSVD in patients with DM.^15^

Adaptation to stress is characterized by an increase in sympathetic activity and a decrease in parasympathetic activity, inducing a state of alertness.^16^ Interestingly, common diseases such as depression, metabolic syndrome, and cancer; smoking habit; and obesity are associated with a decrease in parasympathetic activity and activation of sympathetic activity.^7,17^

One explanation is that DM is a metabolic disease responsible for cardiac autonomic neuropathy, which affects both sympathetic and parasympathetic fibers. DM has a negative influence on almost all HRV parameters, indicating that it leads to cardiac autonomic dysfunction.^18,19^

We demonstrated that an increase in heart rate was associated with higher glucose levels and a decrease in HRV (HF and LF). Although no study has previously assessed this relationship in patients with DM, conflicting results have been reported in the general population, with either high BP associated with an increase in all spectral parameters or a decrease in HRV.^20,21^ It has also been suggested that a decrease in autonomic nervous function precedes the development of clinical hypertension.^22^ However, in our study, there was no significant difference in HRV according to hypertension. Moreover, although age and sex may have a minor role in HRV parameters compared with the variables linked to DM, a previous study demonstrated a decrease in both LF and HF with age and in males.^23,24^ In our study also, HRV was decreased with age and in males.

To the best of our knowledge, this is the first study to simultaneously investigate the HRV and glucose levels in patients with DM using a remote monitoring system. Importantly, in contrast to previous population-based studies,^25,26^ we found that virtually all time- and frequency-domain measures of HRV, either as a composite score or as individual measures, were associated with worsening glucose tolerance. This may be explained by the fact that we used a more accurate 14 days remote-monitoring ECG-derived HRV as opposed to HRV derived from short-term ECG recordings. In addition, we were able to adjust for a large series of potential confounders, including real-time glucose level, respiration, and physical activity, objectively measured in a live studio at our institute using a remote monitoring system.

## Limitations

Our study has some limitations that must be addressed. First, the relatively small sample size was a limiting factor in generalizing the findings to the DM population. However, it was sufficient to identify significant correlations between HRV and glucose levels in individuals with DM using a remote system for HRV and continuous glucose monitoring. Despite the small number of patients, our analysis demonstrated significant and interesting relationships, particularly between the HRV parameters and glucose levels associated with DM. Hence, the results of our study should be considered hypothesis-generating, and future prospective studies are warranted to confirm these results. Second, in the present study, we only evaluated patients with DM aged<75 years. Although a previous study ^27^ in patients with DM and prediabetes and healthy participants and another study ^23^ that investigated the impact of sex and age on HRV demonstrated that HRV indices significantly increased with the participants’ age, we do not know whether older adults with DM aged>75 years have similar or worse HRV patterns than older healthy individuals. Third, the health status of the controls was not detailed in our study, which could have influenced the HRV parameters. This may also have minimized the differences in HRV between patients with DM and controls.

## Conclusions

Poorly controlled glucose levels are independently associated with lower HRV in patients with DM. This was further substantiated by the independent continuous association between real-time measurements of hyperglycemia and lower HRV. These data strongly suggest that cardiac autonomic dysfunction is caused by elevated blood sugar levels.

## Data Availability

all data are available from kosin university gospel hospital (KUGH) Diabetes mellitus registry

## Acknowledgements

We thank all members of the Division of Cardiology, Division Endocrinology, Department of Internal Medicine, Kosin University Gospel Hospital for their assistance and support with data collection. Dr. Chul Ho Oak has received consulting honoraria from Me-zoo and U2meditek and a research grant from Korea Health Industry Development Institute (KHIDI 20210048001). The remaining authors have nothing to disclose.

## Reference

1. Shaffer F, Ginsberg JP. An Overview of Heart Rate Variability Metrics and Norms. Front Public Health. 2017;5:258. doi: 10.3389/fpubh.2017.00258

2. Shah AS, El Ghormli L, Vajravelu ME, Bacha F, Farrell RM, Gidding SS, Levitt Katz LE, Tryggestad JB, White NH, Urbina EM. Heart Rate Variability and Cardiac Autonomic Dysfunction: Prevalence, Risk Factors, and Relationship to Arterial Stiffness in the Treatment Options for Type 2 Diabetes in Adolescents and Youth (TODAY) Study. Diabetes Care. 2019;42:2143–2150. doi: 10.2337/dc19-0993

3. Motairek I, Al-Kindi S. Ameliorating Cardiovascular Risk in Patients with Type 2 Diabetes. Endocrinol Metab Clin North Am. 2023;52:135–147. doi: 10.1016/j.ecl.2022.07.002

4. Sun H, Saeedi P, Karuranga S, Pinkepank M, Ogurtsova K, Duncan BB, Stein C, Basit A, Chan JCN, Mbanya JC, et al. IDF Diabetes Atlas: Global, regional and country-level diabetes prevalence estimates for 2021 and projections for 2045. Diabetes Res Clin Pract. 2022;183:109119. doi: 10.1016/j.diabres.2021.109119

5. Tancredi M, Rosengren A, Svensson AM, Kosiborod M, Pivodic A, Gudbjornsdottir S, Wedel H, Clements M, Dahlqvist S, Lind M. Excess Mortality among Persons with Type 2 Diabetes. N Engl J Med. 2015;373:1720–1732. doi: 10.1056/NEJMoa1504347

6. Sudo SZ, Montagnoli TL, Rocha BS, Santos AD, de Sa MPL, Zapata-Sudo G. Diabetes-Induced Cardiac Autonomic Neuropathy: Impact on Heart Function and Prognosis. Biomedicines. 2022;10. doi: 10.3390/biomedicines10123258

7. Benichou T, Pereira B, Mermillod M, Tauveron I, Pfabigan D, Maqdasy S, Dutheil F. Heart rate variability in type 2 diabetes mellitus: A systematic review and meta-analysis. PLoS One. 2018;13:e0195166. doi: 10.1371/journal.pone.0195166

8. Gerritsen J, Dekker JM, TenVoorde BJ, Kostense PJ, Heine RJ, Bouter LM, Heethaar RM, Stehouwer CD. Impaired autonomic function is associated with increased mortality, especially in subjects with diabetes, hypertension, or a history of cardiovascular disease: the Hoorn Study. Diabetes Care. 2001;24:1793–1798. doi: 10.2337/diacare.24.10.1793

9. Song MH, Cho SP, Kim W, Lee KJ. New real-time heartbeat detection method using the angle of a single-lead electrocardiogram. Comput Biol Med. 2015;59:73–79. doi: 10.1016/j.compbiomed.2015.01.015

10. Deckers JG, Schellevis FG, Fleming DM. WHO diagnostic criteria as a validation tool for the diagnosis of diabetes mellitus: a study in five European countries. Eur J Gen Pract. 2006;12:108–113. doi: 10.1080/13814780600881268

11. Ewing DJ, Boland O, Neilson JM, Cho CG, Clarke BF. Autonomic neuropathy, QT interval lengthening, and unexpected deaths in male diabetic patients. Diabetologia. 1991;34:182–185. doi: 10.1007/BF00418273

12. Verrotti A, Loiacono G, Mohn A, Chiarelli F. New insights in diabetic autonomic neuropathy in children and adolescents. Eur J Endocrinol. 2009;161:811–818. doi: 10.1530/EJE-09-0710

13. Tsuji H, Larson MG, Venditti FJ, Jr., Manders ES, Evans JC, Feldman CL, Levy D. Impact of reduced heart rate variability on risk for cardiac events. The Framingham Heart Study. Circulation. 1996;94:2850–2855. doi: 10.1161/01.cir.94.11.2850

14. Liao D, Carnethon M, Evans GW, Cascio WE, Heiss G. Lower heart rate variability is associated with the development of coronary heart disease in individuals with diabetes: the atherosclerosis risk in communities (ARIC) study. Diabetes. 2002;51:3524–3531. doi: 10.2337/diabetes.51.12.3524

15. Qiu Q, Song W, Zhou X, Yu Z, Wang M, Hao H, Pan D, Luo X. Heart rate variability is associated with cerebral small vessel disease in patients with diabetes. Front Neurol. 2022;13:989064. doi: 10.3389/fneur.2022.989064

16. Hufnagel C, Chambres P, Bertrand PR, Dutheil F. The Need for Objective Measures of Stress in Autism. Front Psychol. 2017;8:64. doi: 10.3389/fpsyg.2017.00064

17. Lin KD, Chang LH, Wu YR, Hsu WH, Kuo CH, Tsai JR, Yu ML, Su WS, Lin IM. Association of depression and parasympathetic activation with glycemic control in type 2 diabetes mellitus. J Diabetes Complications. 2022;36:108264. doi: 10.1016/j.jdiacomp.2022.108264

18. El Tantawy A, Anwar G, Esmail R, Zekri H, Mahmoud S, Samir N, Fathalla A, Maher M, AbdelMassih AF. Cardiac autonomic neuropathy linked to left ventricular dysfunction in type 1 diabetic patients. Cardiovasc Endocrinol Metab. 2022;11:e0272. doi: 10.1097/XCE.0000000000000272

19. Liu L, Wu Q, Yan H, Chen B, Zheng X, Zhou Q. Association between Cardiac Autonomic Neuropathy and Coronary Artery Lesions in Patients with Type 2 Diabetes. Dis Markers. 2020;2020:6659166. doi: 10.1155/2020/6659166

20. Askin L, Cetin M, Turkmen S. Ambulatory blood pressure results and heart rate variability in patients with premature ventricular contractions. Clin Exp Hypertens. 2018;40:251–256. doi: 10.1080/10641963.2017.1356846

21. de Andrade PE, do Amaral JAT, Paiva LDS, Adami F, Raimudo JZ, Valenti VE, Abreu LC, Raimundo RD. Reduction of heart rate variability in hypertensive elderly. Blood Press. 2017;26:350–358. doi: 10.1080/08037051.2017.1354285

22. Schroeder EB, Liao D, Chambless LE, Prineas RJ, Evans GW, Heiss G. Hypertension, blood pressure, and heart rate variability: the Atherosclerosis Risk in Communities (ARIC) study. Hypertension. 2003;42:1106–1111. doi: 10.1161/01.HYP.0000100444.71069.73

23. Reardon M, Malik M. Changes in heart rate variability with age. Pacing Clin Electrophysiol. 1996;19:1863–1866. doi: 10.1111/j.1540-8159.1996.tb03241.x

24. Pavithran P, Madanmohan T, Nandeesha H. Sex differences in short-term heart rate variability in patients with newly diagnosed essential hypertension. J Clin Hypertens (Greenwich). 2008;10:904–910. doi: 10.1111/j.1751-7176.2008.00052.x

25. Ziegler D, Voss A, Rathmann W, Strom A, Perz S, Roden M, Peters A, Meisinger C, Group KS. Increased prevalence of cardiac autonomic dysfunction at different degrees of glucose intolerance in the general population: the KORA S4 survey. Diabetologia. 2015;58:1118–1128. doi: 10.1007/s00125-015-3534-7

26. Meyer ML, Gotman NM, Soliman EZ, Whitsel EA, Arens R, Cai J, Daviglus ML, Denes P, Gonzalez HM, Moreiras J, et al. Association of glucose homeostasis measures with heart rate variability among Hispanic/Latino adults without diabetes: the Hispanic Community Health Study/Study of Latinos (HCHS/SOL). Cardiovasc Diabetol. 2016;15:45. doi: 10.1186/s12933-016-0364-y

27. Coopmans C, Zhou TL, Henry RMA, Heijman J, Schaper NC, Koster A, Schram MT, van der Kallen CJH, Wesselius A, den Engelsman RJA, et al. Both Prediabetes and Type 2 Diabetes Are Associated With Lower Heart Rate Variability: The Maastricht Study. Diabetes Care. 2020;43:1126–1133. doi: 10.2337/dc19-2367

